# Construction of the index system for screening and assessment of dysphagia in Chinese elderly people based on Delphi method

**DOI:** 10.1101/2024.09.19.24314027

**Authors:** Li Gaoqiang, Chen Yue, Yong Qinge

**Affiliations:** Department of Respiratory and Critical Care Medicine, Second Medical Center, PLA General Hospital, 100039; Department of Nursing, Second Medical Center, PLA General Hospital, 100039

**Keywords:** The elderly, Dysphagia, Delphi Technique, Screening, Evaluation

## Abstract

**Background:** Dysphagia is an important factor affecting aspiration pneumonia in the elderly, which will greatly increase the risk of poor prognosis and even death. Early detection, diagnosis and effective prevention are the key to improve the prognosis of patients. However, there is currently no systematic tool for screening and evaluating swallowing disorders in the elderly.

**Objective:** This study aimed to establish an index system for the screening and evaluation of dysphagia in the elderly, and to provide evidence for the screening and evaluation of dysphagia in the community and clinic.

**Methods:** The draft of the index system was determined based on the combination of literature review and clinical practice. The Delphi method was applied to conduct expert correspondence consultation, and the index system for screening and evaluation of dysphagia in the elderly was established. The weight of each index was determined by analytic hierarchy process.

**Results:** A total of 19 experts in related fields were consulted for 3 rounds. The questionnaire recovery rates were 94.7%, 100% and 100%, respectively. 17 (89.5%), 14 (73.7%) and 5 (26.3%) experts put forward modification opinions, respectively. The expert authority coefficient was 0.920, and the Kendall harmony coefficient was 0.219, 0.261 and 0.306, respectively, with statistical significance (P < 0.001). Finally, the index system for the screening and evaluation of dysphagia in the elderly includes 3 first-level indicators, 10 second-level indicators and 26 third-level indicators.

**Conclusion:** The experts in this study are highly motivated and authoritative, and the established index system for the screening and evaluation of dysphagia in the elderly is scientific, reasonable and targeted, which can provide reference for the screening and evaluation of dysphagia in the community and clinical elderly patients.

## 1. Introduction

With the accelerated aging of Chinese society, the number of elderly population is increasing, and it is expected to reach 487 million by 2050 [1]. Therefore, the society has paid more and more attention to the problems of elderly population. Due to advanced age and various diseases (such as Parkinson’s disease and stroke), elderly patients often develop degenerative changes in sensory-motor physiology related to swallowing function, eventually leading to swallowing disorders [2]. Swallowing disorder is a kind of elderly syndrome, and foreign studies have found that 10%-33% of elderly people are affected by swallowing disorder [3]. According to a meta-analysis in China, the prevalence rate of dysphagia among the elderly in China is as high as 66% and increases with age [4]. Once dysphagia occurs in the elderly and is not detected and treated in time, it is easy to have complications such as aspiration, aspiration pneumonia and malnutrition, which will greatly increase the risk of poor prognosis and even death of patients [5].

Early screening and assessment of swallowing function and implementation of reasonable dietary intervention are considered to be standard treatment for swallowing disorders [6]. However, most elderly patients are not aware of their own swallowing dysfunction, and do not think that coughing or aspiration is the manifestation of swallowing disorder in the elderly [7]. Therefore, it is particularly necessary to carry out comprehensive and systematic screening and evaluation of swallowing disorders in the elderly. Currently, commonly used screening tools for swallowing disorders include Eating Assessment Questionnaire-10 (EAT-10) [8], Kota drinking water test [9], volume-viscosity swallowing test (V-VST) [10], etc. EAT-10 lacks objectivity as a single screening tool despite its wide application range, short time consuming and simple operation method. Although the Lowada drinking water test is characterized by objectivity and high sensitivity, its specificity is poor (20%-30%) [11]. V-VST can test the ability of elderly patients to swallow food and promote the eating plan for the elderly based on the screening results, but the swallowing test starts from the swallowing semi-solid ability and ignores the swallowing water ability. Studies have shown that the risk of aspiration when swallowing water in patients with swallowing disorders is much higher than that of swallowing semi-solid [12,13]. Although these evaluation tools have the advantages of low cost, easy operation and short time consuming, the screening and evaluation effect of a single tool is poor due to poor tolerance of the elderly, not obvious symptoms of swallowing disorders, many underlying diseases and poor coordination [14]. At present, there is no evaluation system for the screening and evaluation of dysphagia in the elderly, and it is impossible to systematically and scientifically screen and evaluate dysphagia in elderly patients. Therefore, this study establishes a scientific and systematic index system for the screening and evaluation of dysphagia in the community and clinical elderly through literature research and expert consultation. To provide evidence for screening and evaluation of dysphagia in elderly patients.

## 2. Material and methods

### 2.1 Establish a research group

Team members include a director of nursing department (chief nurse), mainly responsible for the overall research design and quality control; A chief nurse (chief nurse) is mainly responsible for the formulation of specific items in the first draft and the selection of experts; Three nurses (Master of Nursing) are mainly responsible for literature research, compilation, distribution and recovery of consultation questionnaires, modification of consultation opinions, and data analysis and sorting.

### 2.2 The first draft of the index system for screening and evaluation of dysphagia in the elderly was formed

In “the elderly/elderly patients”, “dysphagia/dyspharyngeal” “screening/evaluation/management” and “expert consensus/summary of “evidence/ system review /practice guidelines/manual” was used to search Chinese databases such as CNKI, Wanfang Database, China Biomedical Literature Database, Yimaitong, English databases such as PubMed, Embase, UpToDate and BMJ best practice, CINAHL, Cochrane Library. The article information is extracted by checking, screening and evaluating the retrieved literature. Through the joint discussion of team members and clinical experts, a draft of the screening and evaluation index system for swallowing disorders in the elderly was formed, including 3 items of class I, 10 items of class II, and 23 items of class III.

### 2.3 Develop expert consultation questionnaire

The expert consultation questionnaire mainly includes the following contents: (1) Introduction of the questionnaire: explain the content, purpose and significance of the research; (2) Consultation table for the first draft of the index system for screening and evaluation of dysphagia in the elderly. Each expert can make a judgment of “agree/disagree” according to his or her own experience, and at the same time give modification suggestions for each item. The importance evaluation of indicators at each level adopts Likert 5-level scoring method, that is, 1-5 points respectively indicate “very unimportant ∼ very important”; ➂ The questionnaire of the basic information of the experts, including gender, age, work unit and department, title, education, professional occupation, working years, etc.; ➃ Authority level questionnaire: the expert’s familiarity with the content filled in the questionnaire and the influence of the judgment basis on the expert’s judgment. The familiarity is assigned 1.0, 0.8, 0.5, 0.2 and 0 points according to “very familiar, relatively familiar, generally familiar, not very familiar, not familiar”; The judgment basis is divided into four dimensions, all of which are quantified by “large, medium and small”. The dimensions of “theoretical knowledge” are assigned 0.3, 0.2 and 0.1 points respectively, the dimensions of “practice or scientific research experience” are assigned 0.5, 0.4 and 0.3 points respectively, and the “reference to domestic and foreign literatures” and “intuitive feeling of experts” are assigned 0.1 points respectively.

### 2.4 Selection of experts

Inclusion criteria: ➀ Medical experts, nursing experts and nursing management experts with certain academic level in the field of swallowing disorders in the elderly, familiar with the field of screening and evaluation of swallowing disorders at home and abroad; ➁ Associate senior title or above, more than 10 years of work experience; ➂ Agree to participate in this study and be able to complete expert consulting work.

### 2.5 Implementation expert consultation

From April to August 2023, questionnaires were distributed and returned by paper and email. Each round of expert opinions was returned within 2 weeks. After each round of questionnaire collection, members of the research team sorted and summarized the questionnaires, combined with the entry screening criteria and expert feedback, the items were added or modified, and the next consultation questionnaire was formed and issued. Until the third round of expert consultation, all expert opinions basically reached a consensus, that is, the consultation was stopped. The inclusion criteria of the articles were the mean value of importance assignment > 3.5 points and the coefficient of variation < 0.25[15].

### 2.6 Statistical analysis

Excel 2021, SPSS20.0 and Yaahp 12.0 were used for data entry and analysis. The basic situation of consulting experts is described by frequency and percentage, and the positive degree of experts is reflected by the questionnaire recovery rate and the number of opinions submitted. The expert’s authority coefficient is calculated using the arithmetic average of the familiarity coefficient and the judgment coefficient. The degree of expert opinion coordination is expressed by Kendall harmony coefficient and variation coefficient. The software Yaahp 12.0 was used to determine the weight of each index, and the test level α=0.05.

## 3 Results

### 3.1 Basic information about consulting experts

A total of 19 experts from geriatric medicine, geriatric rehabilitation, geriatric gastroenterology, geriatric neurology and other departments participated in 3 rounds of expert consultation. Among them, 8 were males and 11 were females, aged 37-65 (50.89 ±8.10) years old, 4 were 11-20 years old, 6 were 21-30 years old, and 9 were over 30 years old. The research fields include geriatric clinical medicine 8 people, geriatric nursing 7 people, nursing management 2 people, scientific research and teaching 2 people; Professional titles for senior 10, deputy senior 9; The academic qualifications are 9 doctor’s degrees, 6 master’s degrees and 4 bachelor’s degrees.

### 3.2 The degree of motivation and authority of the expert

19 experts were consulted, and the recovery rates of the three rounds of consultation questionnaires were 94.7%, 100% and 100%, respectively. Among them, 17 (89.5%), 14 (73.7%) and 5 (26.3%) experts put forward suggestions for revision, and the experts were more active. The coefficient of expert familiarity is 0.86, the coefficient of judging basis is 0.98, and the coefficient of expert authority is 0.92.

### 3.3 Degree of coordination of expert opinions

According to the results of **Table 1**, the Kendall harmony coefficients of the three rounds of consultation expert opinions are 0.219, 0.261, and 0.306 respectively (P < 0.001), indicating that expert opinions gradually converge.

**Table 1.** Degree of coordination of expert opinions.

| items | First round of consultation |  |  | Second round of consultation |  |  | Third round of consultation |  |  |
| --- | --- | --- | --- | --- | --- | --- | --- | --- | --- |
|  | Kendall coefficient of concordance | Chi square value | P-value | Kendall coefficient of concordance | Chi square value | P-value | Kendall coefficient of concordance | Chi square value | P-value |
| Primary index | 0.389 | 14.774 | <0.001 | 0.419 | 15.935 | <0.001 | 0.471 | 17.882 | <0.001 |
| Secondary index | 0.284 | 48.597 | <0.001 | 0.302 | 51.571 | <0.001 | 0.398 | 68.101 | <0.001 |
| Three-level index | 0.192 | 91.041 | <0.001 | 0.246 | 116.839 | <0.001 | 0.271 | 128.506 | <0.001 |
| totality | 0.219 | 157.934 | <0.001 | 0.261 | 188.508 | <0.001 | 0.306 | 220.797 | <0.001 |

### 3.4 Expert consultation results

After 3 rounds of expert consultation, according to the indicator screening criteria, combined with the opinions of experts and the discussion of team members, some indicators have been added, subtraction and modified, and the revisions are as follows: ➀ Modify the “evaluation strategy logic” in the first-level indicator to “evaluation strategy logic between the above two”. Experts believe that this index system is divided into two parts: rapid screening and accurate evaluation, and there is a logical connection between the two parts. Therefore, it should be clearly pointed out that the object of evaluation strategy logic is between the above two (rapid screening and accurate evaluation). ➁ In the secondary index, the “all items in rapid screening are normal, then the evaluation is terminated” is modified to “all items in rapid screening are normal, then regular assessment (evaluation every three months) or immediate assessment when there are changes (such as coughing, etc.)”, experts believe that for elderly patients, especially elderly people, their conditions change rapidly. The onset of dysphagia can occur within a few months, so for older people who have normal entries in rapid screening, regular evaluation is needed to detect dysphagia in a timely manner; Secondly, swallowing disorder is an insidious symptom that requires medical staff to immediately evaluate elderly patients when they have changes (such as coughing when eating water), and the whole screening and evaluation system reflects continuity and dynamics. ➂ Change the “medical history screening” in the secondary index to “medical history inquiry related to swallowing disorders”, experts believe that “medical history screening” is too broad and not targeted, while “medical history inquiry related to swallowing disorders” is more specific, clear, and closely related to the research theme. (4) The entry “swallowing functional exchange assessment” in rapid screening is moved to the entry of accurate assessment. Experts believe that the “swallowing functional exchange assessment” consists of 7 items, which requires experienced medical staff to ask the patient’s eating situation in detail in order to obtain the rating. The process is more complicated and is not suitable for inclusion in the rapid screening entry. ➄ In the “dysphagia related medical history” entry corresponding to the tertiary indicators added “clear history of aspiration pneumonia”, “oropharyngeal diseases” and “hoarseness after swallowing” 3 entries, experts believe that aspiration pneumonia is often caused by aspiration in elderly patients, and in the reason for aspiration, swallowing disorders accounted for a large proportion, so it needs to be taken into account. Secondly, the presence of oropharyngeal diseases is likely to affect the swallowing function of patients, resulting in swallowing disorders, so it is recommended to add. At the same time, the addition of “hoarseness after swallowing” is to take into account that the patient’s swallowing disorder causes food to enter the trachea, which may lead to changes in the voice. The final screening and evaluation index system for dysphagia in the elderly includes 3 first-level indicators, 10 second-level indicators and 26 third-level indicators, the contents of which are shown in Table 2.

**Table 2.**
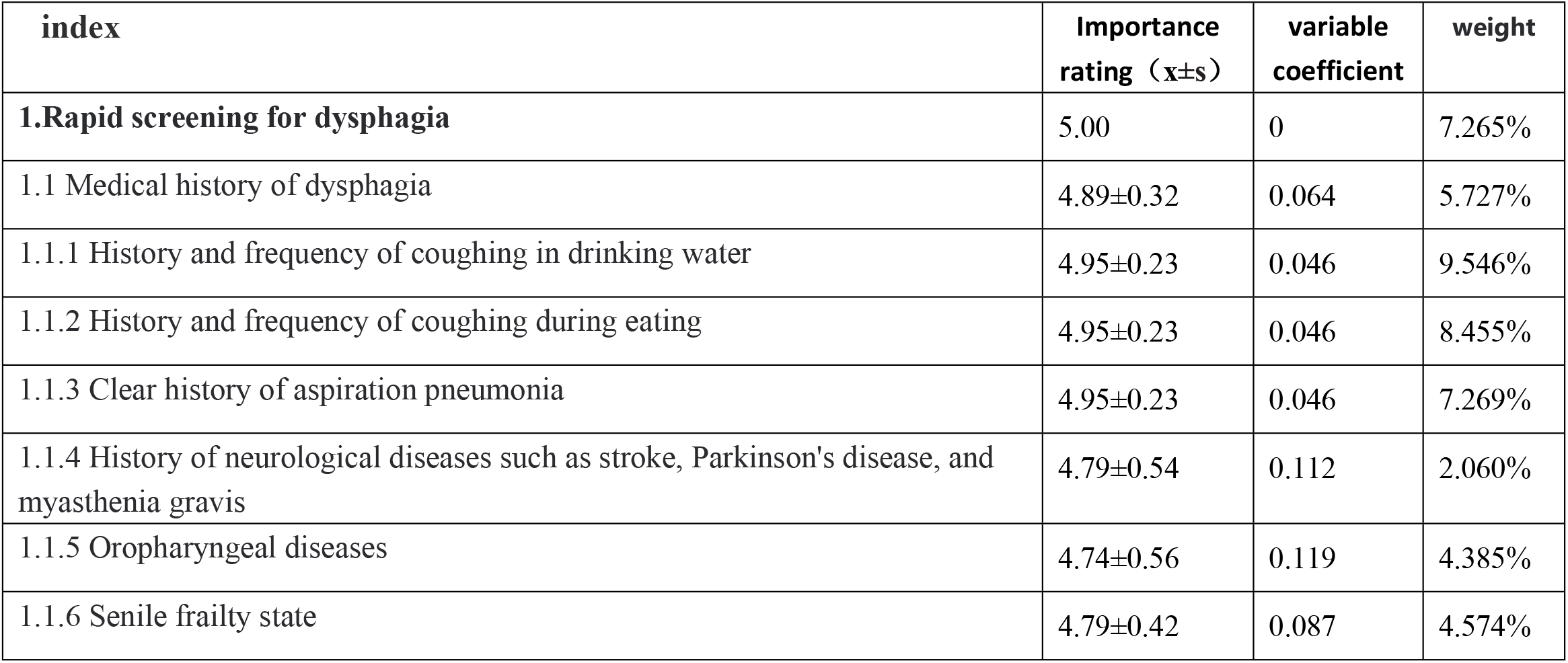
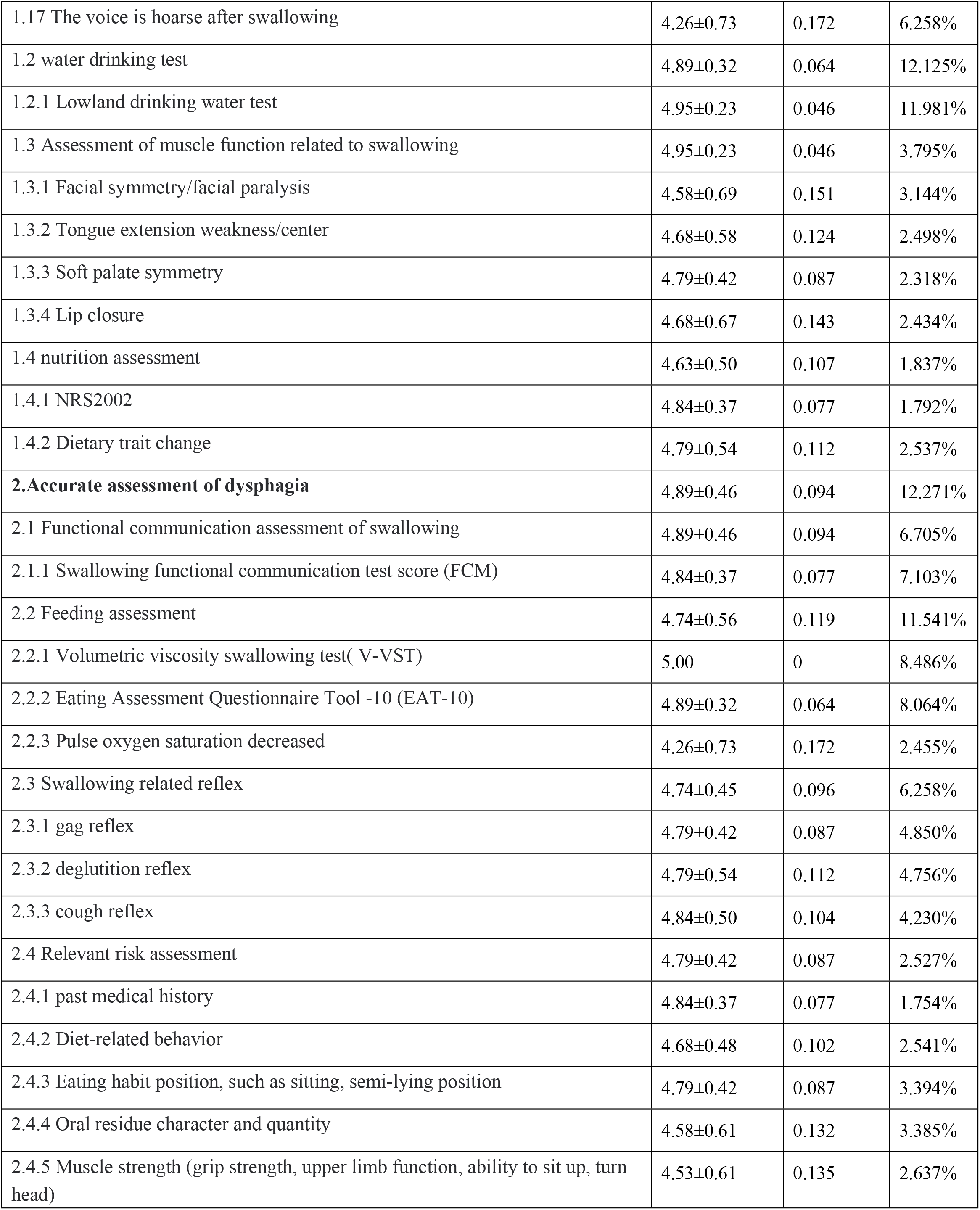
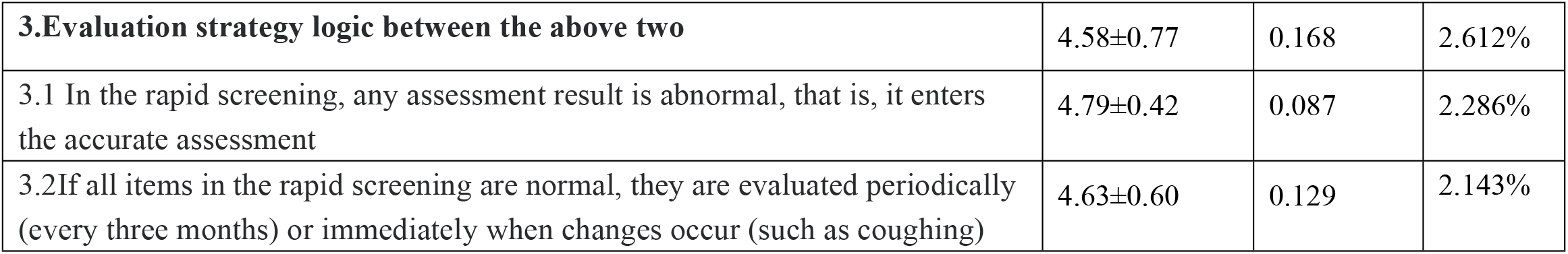
Screening and evaluation index system for dysphagia in the elderly.

| index | Importance rating ($\bar{x} \pm s$) | variable coefficient | weight |
| --- | --- | --- | --- |
| **1.Rapid screening for dysphagia** | 5.00 | 0 | 7.265% |
| 1.1 Medical history of dysphagia | 4.89±0.32 | 0.064 | 5.727% |
| 1.1.1 History and frequency of coughing in drinking water | 4.95±0.23 | 0.046 | 9.546% |
| 1.1.2 History and frequency of coughing during eating | 4.95±0.23 | 0.046 | 8.455% |
| 1.1.3 Clear history of aspiration pneumonia | 4.95±0.23 | 0.046 | 7.269% |
| 1.1.4 History of neurological diseases such as stroke, Parkinson's disease, and myasthenia gravis | 4.79±0.54 | 0.112 | 2.060% |
| 1.1.5 Oropharyngeal diseases | 4.74±0.56 | 0.119 | 4.385% |
| 1.1.6 Senile frailty state | 4.79±0.42 | 0.087 | 4.574% |
| 1.17 The voice is hoarse after swallowing | 4.26±0.73 | 0.172 | 6.258% |
| 1.2 water drinking test | 4.89±0.32 | 0.064 | 12.125% |
| 1.2.1 Lowland drinking water test | 4.95±0.23 | 0.046 | 11.981% |
| 1.3 Assessment of muscle function related to swallowing | 4.95±0.23 | 0.046 | 3.795% |
| 1.3.1 Facial symmetry/facial paralysis | 4.58±0.69 | 0.151 | 3.144% |
| 1.3.2 Tongue extension weakness/center | 4.68±0.58 | 0.124 | 2.498% |
| 1.3.3 Soft palate symmetry | 4.79±0.42 | 0.087 | 2.318% |
| 1.3.4 Lip closure | 4.68±0.67 | 0.143 | 2.434% |
| 1.4 nutrition assessment | 4.63±0.50 | 0.107 | 1.837% |
| 1.4.1 NRS2002 | 4.84±0.37 | 0.077 | 1.792% |
| 1.4.2 Dietary trait change | 4.79±0.54 | 0.112 | 2.537% |
| **2.Accurate assessment of dysphagia** | 4.89±0.46 | 0.094 | 12.271% |
| 2.1 Functional communication assessment of swallowing | 4.89±0.46 | 0.094 | 6.705% |
| 2.1.1 Swallowing functional communication test score (FCM) | 4.84±0.37 | 0.077 | 7.103% |
| 2.2 Feeding assessment | 4.74±0.56 | 0.119 | 11.541% |
| 2.2.1 Volumetric viscosity swallowing test( V-VST) | 5.00 | 0 | 8.486% |
| 2.2.2 Eating Assessment Questionnaire Tool -10 (EAT-10) | 4.89±0.32 | 0.064 | 8.064% |
| 2.2.3 Pulse oxygen saturation decreased | 4.26±0.73 | 0.172 | 2.455% |
| 2.3 Swallowing related reflex | 4.74±0.45 | 0.096 | 6.258% |
| 2.3.1 gag reflex | 4.79±0.42 | 0.087 | 4.850% |
| 2.3.2 deglutition reflex | 4.79±0.54 | 0.112 | 4.756% |
| 2.3.3 cough reflex | 4.84±0.50 | 0.104 | 4.230% |
| 2.4 Relevant risk assessment | 4.79±0.42 | 0.087 | 2.527% |
| 2.4.1 past medical history | 4.84±0.37 | 0.077 | 1.754% |
| 2.4.2 Diet-related behavior | 4.68±0.48 | 0.102 | 2.541% |
| 2.4.3 Eating habit position, such as sitting, semi-lying position | 4.79±0.42 | 0.087 | 3.394% |
| 2.4.4 Oral residue character and quantity | 4.58±0.61 | 0.132 | 3.385% |
| 2.4.5 Muscle strength (grip strength, upper limb function, ability to sit up, turn head) | 4.53±0.61 | 0.135 | 2.637% |
| **3.Evaluation strategy logic between the above two** | 4.58±0.77 | 0.168 | 2.612% |
| 3.1 In the rapid screening, any assessment result is abnormal, that is, it enters the accurate assessment | 4.79±0.42 | 0.087 | 2.286% |
| 3.2 If all items in the rapid screening are normal, they are evaluated periodically (every three months) or immediately when changes occur (such as coughing) | 4.63±0.60 | 0.129 | 2.143% |

### 3.5 Establishment of screening and evaluation index system weight for dysphagia in the elderly

Based on the importance assigned by the experts to each item of the screening and evaluation indicators for swallowing disorders in the elderly in the third round of expert consultation, the judgment matrix was constructed with reference to the Saaty1-9 scale method, and the data was input into Yaahp 12.0 software to calculate the weight coefficients of each level of indicators. The consistency test results (CR value) of all levels of indicators were <0.1, as shown in **Table 2**.

## 4. Discussion

Based on the research team’s full search and summary of domestic and foreign literature, this study established a preliminary indicator system in combination with the actual clinical situation, added, deleted and modified the indicators through three rounds of Delphi expert consultation, and quantified the weight of each indicator by analytic hierarchy process, forming the final screening and evaluation indicator system for dysphagia in the elderly, which is of good scientific nature. The reliability of research results is related to the source and authority of experts, their enthusiasm for consultation and the degree of coordination of experts [16]. The 19 experts selected in this study are all engaged in the treatment, nursing, rehabilitation, scientific research and management of elderly patients with more than 10 years of work, and have rich clinical experience. Among them, 10 experts are senior and 9 are deputy senior. There are 9 doctoral degrees, 6 master degrees and 4 bachelor degrees, which can provide guidance in the process of elderly dysphagia screening and evaluation indicators. In the three rounds of expert consultation, the recovery rates of the consultation questionnaires were 94.7%, 100% and 100% respectively, among which the opinion submission rates were 89.5%, 73.7% and 26.3% respectively, indicating that the experts had a high enthusiasm. The expert authority coefficient represents the quantitative index of the authority degree of experts in this research field. Cr > 0.7 indicates that experts have a good degree of trust and have a greater grasp of this field [17]. The expert authority coefficient of this study is 0.92, indicating that experts have high authority in the field of swallowing disorders in the elderly, which ensures the credibility of the results of this study. Kendall harmony coefficients of the three rounds of expert consultation were 0.219, 0.261 and 0.306 respectively (P < 0.001), indicating that the experts’ evaluation opinions on the indicators gradually converged, and the above indicators showed that the reliability of the research results was guaranteed from the expert level. In terms of content analysis, at the beginning of the construction of this study, reference was made to relevant domestic and international guidelines or expert consensus [18-19]. In the screening part, medical history inquiry, drinking water test, physical examination, nutritional screening and other contents were included, and the first draft was written in combination with clinical practice to improve the scientificity and feasibility of the first draft.

The consensus of Chinese experts points out that early screening, assessment and targeted management of swallowing function in elderly patients can effectively prevent aspiration, reduce the incidence of aspiration pneumonia, shorten the length of hospital stay and improve patient satisfaction [20]. Based on this, this study took elderly patients as the research object, fully considered the physiological characteristics of elderly people prone to frailty [21], and developed a targeted and convenient elderly dysphagia screening and evaluation index system. First of all, in order to quickly screen the elderly population with swallowing disorders, this index system sets two stages of rapid screening and accurate assessment. In the rapid screening stage, simple medical history inquiry and drinking water test are carried out. If there is at least one positive result, accurate assessment will be entered. On the one hand, rapid screening reduces the scope of screening and saves a lot of labor and time costs; On the other hand, the rapid screening of the problem is followed by a detailed evaluation of the swallowing disorder, taking into account the continuity between screening and evaluation. Through rapid screening of each elderly patient, patients with positive results were selected, and then accurate assessment was carried out to clarify the influential factors affecting the elderly’s swallowing function, providing a reference for targeted intervention.

## 5. Conclusions

Through literature research and 3 rounds of Delphi expert consultation, this study finally established a screening and evaluation index system for dysphagia in the elderly, including 3 first-level indicators, 10 second-level indicators and 26 third-level indicators. The content has scientific and clinical practical value, and can provide guidance for the screening and evaluation of dysphagia in the elderly. The operability, sensitivity and specificity of the index system will be further verified in the future.

## Data Availability

All relevant data are within the manuscript and its Supporting Information files.

## References

[1] [Chen Y M, Liu Z F, Li X D, et al. Aging trend and elderly population forecast in China from 2015 to 2050 [J]. Chinese Journal of Social Medicine, 2018, 35(05):480–483.

[2] Logemann JA, Curro FA, Pauloski B, et al. Aging effects on oropharyngeal swallow and the role of dental care in oropharyngeal dysphagia. Oral Dis. 2013; 19 (8): 733–737.

[3] Thiyagalingam S, Kulinski AE, Thorsteinsdottir B, et al. Dysphagia in Older Adults. Mayo Clin Proc. 2021 Feb; 96 (2): 488–497.

[4] Liu Yaxin, Jiang Yunlan, Huang Xiaoxing et al. Meta-analysis of the prevalence of dysphagia in elderly Chinese [J]. Chinese Journal of General Medicine, 2019, 26(12):1496-1502+1512. (in Chinese)

[5] Swan K, Speyer R, Heijnen BJ, et al. Living with oropharyngeal dysphagia: effects of bolus modification on health-related quality of life--a systematic review. Qual Life Res. 2015 Oct; 24 (10): 2447–56.

[6] Garcia JM, Chambers E 4th. Managing dysphagia through diet modifications. Am J Nurs. 2010 Nov; 110 (11): 26–33; quiz 34-5.

[7] Cao Yanju, Feng Ruijuan, Niu Lijun et al. Analysis of prevalence and related factors of dysphagia among military retired elderly in Beijing [J]. Chinese Journal of Geriatric Cardio-Cerebrovascular Diseases, 2019, 25(03):286–288. (in Chinese)

[8] Belafsky PC, Mouadeb DA, Rees CJ, et al. Validity and reliability of the Eating Assessment Tool (EAT-10). Ann Otol Rhinol Laryngol. 2008; 117 (12): 919–924.

[9] Osawa A, Maeshima S, Tanahashi N. Water-swallowing test: screening for aspiration in stroke patients. Cerebrovasc Dis. 2013; 35 (3): 276–281.

[10] Clave P, Arreola V, Romea M, et al. Accuracy of the volume-viscosity swallow test for clinical screening of oropharyngeal dysphagia and aspiration. Clin Nutr. 2008; 27 (6): 806–815.

[11] Kertscher B, Speyer R, Palmieri M,et al. Bedside screening to detect oropharyngeal dysphagia in patients with neurological disorders: an updated systematic review. Dysphagia. 2014; 29 (2): 204–212.

[12] John JS, Berger L. Using the gugging swallowing screen (GUSS) for dysphagia screening in acute stroke patients. J Contin Educ Nurs. 2015; 46 (3): 103–104.

[13] Li Hui, Feng Hui, Chen Huijing, et al. Application of dysphagia screening tool in elderly care services [J]. Chinese Journal of Rehabilitation Medicine, 2019, 35(03):356–360. (in Chinese)

[14] Tian Xueying, Sun Shu, Liu Siqi, et al. Application of dysphagia screening tool in elderly people in community [J]. Evidence-based Nursing, 2019, 9(01):67–70. (in Chinese)

[15] LV Mengju, Liu Junjie, Li Xuelin. Construction of diet management scheme for patients with dysphagia [J]. Chinese Journal of Nursing, 202, 57(12):1427–1434. (in Chinese)

[16] Tan Xin. Establishment of evidence-based management program and effect evaluation for stroke patients with dysphagia [J]. China Health Care Industry, 2019, 17(19):40–42. (in Chinese)

[17] Cia? kowska M, Adamowski T, Piotrowski P, et al. Czym jest metoda Delphi? Zalety i ograniczenia [What is the Delphi method? Strengths and shortcomings]. Psychiatr Pol. 2008; 42 (1): 5–15.

[18] Yang S, Park JW, Min K, et al. Clinical Practice Guidelines for Oropharyngeal Dysphagia. Ann Rehabil Med. 2023; 47(Suppl 1):S1–S26.

[19] Expert Consensus Group on Rehabilitation Assessment and Treatment of swallowing disorders in China. Expert Consensus on the Evaluation and Treatment of swallowing disorders in China (2017 edition) Part I evaluation [J]. Chinese Journal of Physical Medicine and Rehabilitation, 2017, 39(12):12.

[20] Nutrition and Food Safety Branch of Chinese Geriatric Society, China Center for Evidence-Based Medicine, Editorial Board of Chinese Journal of Evidence-Based Medicine, etc. Chinese expert Consensus on Family nutrition Management in elderly patients with dysphagia (2018 edition)[J]. Chinese Journal of Evidence-Based Medicine, 2018, 18(06):547–559.

[21] Tao Yang, Guo Honghua, Yi Huanying, et al. Analysis of the correlation between oral weakness and body weakness in the elderly [J]. China Chronic Disease Prevention and Control, 2019, 30(12):957–960.

